# The effects of physical distancing and lockdown to restrain SARS-CoV-2 outbreak in the Italian Municipality of Cogne

**DOI:** 10.1101/2021.03.19.21253962

**Authors:** F. Truc, G. Gervino

**Affiliations:** Dipartimento di Fisica, Università di Torino, via P. Giuria 1, 10125 Torino (Italy)

## Abstract

The outbreak of SARS-CoV-2 started in Wuhan, China, and is now a pandemic. An understanding of the prevalence and contagiousness of the disease, and of whether the strategies used to contain it to date have been successful, is important for understanding future containment strategies. One strategy for controlling the spread of SARS-CoV-2 is to adopt strong social distancing policies. The Municipality of Cogne (I), adopted strict lockdown rules from March 4, 2020 up to May 18, 2020. This first wave of the pandemic impressed by the extremely low impact of the SARS-CoV-2 on the locals, compared to the number accused on all the Italian territory. Starting from October 2020 up to the end of December, when the second wave hit Italy and Cogne territory, heavier effects were observed. In order to cast light on the effectiveness of the adopted strategy 74,5% of the local population underwent to a blood screening to detect IgM and IgG antibodies and after six months all the people tested positive were again investigated to establish the longitudinal changes in antibodies level. Moreover, within the context of this survey a rare and interesting case of secondary infection has been identified and here presented.

## 1. Introduction

The ongoing coronavirus disease, designated COVID-19, is caused by the positive-strand RNA virus SARS-CoV-2 and emerged in the Chinese city of Wuhan at the end of 2019. Virus quickly spread to virtually every corner of the globe. Among European countries, Italy is the first and one of the hardest hit by the SARS-CoV-2 pandemic (with more than 2.700.000 confirmed cases and 90.000 deaths by February 15, 2021). In this paper we present and discuss the spread of the pandemic and the adopted strategies to contain it in the Municipality of Cogne, a renowned national and international Italian tourist destination for skiing and excursions, located in the Aosta Valley at 1544 meters of altitude with 1369 inhabitants and an urban density of 1080 persons per km2. Cogne is at the top of a lateral valley it gave its name to, at the bottom of the Gran Paradiso massif (North-West of Italian Alps): a “cul de sac” with a single access route, a unique situation to study the effect of the pandemic. On the weekend February 28-29, 2020, a super-spreading event happened: the massive influx of more than 2000 skiers in Cogne. It was followed on March 4, 2020 by the shutdown of the Municipality established by the local authority, few days before the national lockdown set on March 9, to prevent and limit the spread of the epidemic on all Italian territory. Strong social distancing policies included border-control, closures of schools, shops, hotels, restaurants and services excluded essential businesses, but despite these strict precautions few days later, on March 13, it occurs the first confirmed SARS-CoV-2 symptomatic infection among Cogne residents. On May 25, 2020 the total number of inhabitants diagnosed SARS-CoV-2 positive, since March 4, resulted surprisingly low with respect to the Italian national situation: five only. In a strangely similar way during the Great Influenza Pandemic of 1918 (unfairly known as the Spanish Flu) when the mortality rate for the H1N1 virus in Italy was on average 2,8%, Cogne, 1440 inhabitants on 1918-19, counted only three deaths attributable to Spanish flu epidemic [1]. On 4 and 5 June 2020 Cogne’s Municipality, impressed by the extremely low impact of the SARS-CoV-2 on the locals, launched and funded a population-based serological assay with the following purposes:

– to quantify the seropositivity, detected via the determination of SARS-CoV-2 antibodies, in residents,
– to assess the changes in antibodies levels within 6 months after the first sample collection,
– to study the spread of the virus and the transmission dynamics through the whole territory of Cogne’s Municipality.

A population-based assay, unlike the epidemiological monitoring of ascertained COVID-19 cases, is a survey that is carried out to a representative population sample.Whether that target population is a country, a state or some other group (Cogne population in this case), the population depicted by the sample should be representative of the whole group to which the researcher aims to extend his investigation. 74.5% of the Cogne population were enrolled in the present study (a very high percent indeed), all people under 14 years old were excluded and the age and sex distribution of the sample strictly reflects the entire Cogne population as is is shown in Tab.1. Here we describe and summarize the findings of the above study, conducted between March and December 2020, with the aim of exploring the landscape of an isolate population immunity and its susceptibility to the SARS-CoV-2.

**Tab 1.** In the first column the composition of the June 4-5 immunoassay sample is shown by sex and age bin: notice the very good percent agreement with the composition of Cogne eligible resident population (the last column). Children under 14 years old and Cogne’s residents actually domiciled in other places are excluded from the present study.

| Age composition |  |  |  |  |
| --- | --- | --- | --- | --- |
|  | Participants in the<br>4/5 june<br>immunoassay | % | Cogne's eligible<br>study population | % |
| OVERALL | 857 | 74.5% | 1149 | 100% |
| SEX |  |  |  |  |
| male | 393 | 45.8% | 538 | 46.8% |
| female | 464 | 54.1% | 611 | 53.2% |
| AGE-YEARS |  |  |  |  |
| 14-24 | 130 | 15.1% | 156 | 13,6% |
| 25-49 | 261 | 30.4% | 379 | 32.9% |
| 50-75 | 357 | 41.6% | 463 | 40.3% |
| >= 76 | 109 | 12.7% | 151 | 13.1% |

## 2. Methods

### 2.1 Sample collection

Cogne’s residents, on 4 and 5 June 2020 were invited by the municipal authority to donate a blood sample for subsequent laboratory test to detect antibodies against SARS-CoV-2.

At three designed sampling points were collected blood samples from 857 study participants, corresponding to 74,5% of the eligible (1149) study population, i.e. people living to Cogne down town and spending there a considerable fraction of their time. Children under 14 years old were excluded from the survey. Blood samples were labelled, stored refrigerated and delivered to the laboratory Alliance Medical -Istituto Salus in Genova, for the analysis, at the end of each of the above two days. On December 15, 2020 (second time-point) Cogne’s Municipality invited people tested positive for IgM and/or IgG [2],[3] to the second blood test, to assess the longitudinal changes in antibodies level within 6 months after the first sample collection. For each participant were collected information on sex, age and home address. All study participants provided written informed consent. The study was approved by the Ethics Committee of Aosta Valley (I).

### 2.2 Antibody measurement

The serological test was a chemiluminescent micro-particle immunoassay (YHLO iFlash; Pantec s.r.l.) for quantitative detection of IgG (C86095G) and IgM (C86095M) against SARS-CoV-2 Nucleoprotein (N) and Spike glycoprotein (S) [4]. The manufacturer supplied thresholds for positivity and reported the following data:

sensitivity (IgG) 97.3%, sensitivity (IgM) 86.1%,

specificity (IgG) 96.3%, specificity (IgM) 99.2%.

### 2.3 Nasopharyngeal swabs

Testing by quantitative polymerase-chain-reaction (qPCR) [5] has become widespread in Italy with the purpose of suppress outbreaks and controlling the transmission and viral spread and more than 33.000.000 qPCR tests were done up to February 15, 2021 on all the Italian territory. Since the beginning of the pandemic Cogne’s inhabitants too went repeatedly under to nasopharyngeal swab tests for the detection of SARS-CoV-2 by the Prevention Department of the Regional Health Service. After a starting period of double testing at the Italian National Institute of Health, the Regional Reference Laboratory (Laboratorio di Analisi dell’Azienda U.S.L. della Valle d’Aosta) received accreditation as Reference Laboratory of Covid-19 testing.

### 2.4 Funding sources

The project was budgeted under the contract COGNE-COVID by the Municipality of Cogne. The funder granted access at acquisition of the results of serological tests and nasopharyngeal swabs carried out in Cogne between March and December 2020, but played no part in the design, data analysis and writing. The two authors are responsible for the decision to submit for publication.

## 3. Results

### 3.1 First wave

Between March 4 and June 4, 2020 in the municipality of Cogne were collected 107 nasopharyngeal swabs and identified 5 positive cases. All the people (4 men, 1 female) tested positive resulted symptomatic. (We adopt the following definition of Covid-19 symptomatic i.e. a person who reports fever above 37°C and/or at least two of the following symptoms: headache, conjunctivitis, anosmia and/or ageusia, sore throat, diarrhea, asthenia, shortness of breath, muscle and/or joint pain.). The 5 patients, confirmed symptomatic by treating physicians, were diagnosed qPCR-positive at the following dates: March 13, 14, 15, 17, 22, 2020. Nobody required hospitalization. No other symptomatic and/or qPCR positive cases were identified until June 4, 2020, the start date of the serological test, and this was the situation up to October 2020. All symptomatic cases were found to belong to a single flare-up developed in Municipal House, where the few essential businesses excluded by the lockdown were centered. On June 4-5, 2020 SARS-CoV-2 antibodies (IgM, IgG) of up to 857 residents (74.5% of eligible individuals) were measured, via serological immunoassay, all the 5 people positive to nasopharyngeal swabs underwent to serological test as well. The serological test identified 29 people IgG positives and, among them, 4 IgM positives (see Tab.2). On this first survey the seroprevalence was 3,38% and this is the incidence of persons infected. Between the proved infected the asymptomatic cases were 82,8%. It is worth to notice that between the proved infected 4 out of 29 (13.8%) showed the presence of both antibodies (IgM and IgG). All symptomatic people resulted positive to both nasopharyngeal swabs and IgG antibodies and negative to IgM antibodies. The remaining 24 people IgG positive (4 of them also IgM positive) were totally asymptomatic. In Tab.2 the results of the screening of June 2020 are shown. Incidence of the illness is equivalent between females and males and all symptomatic people are concentrated in the age interval between 50 and 74 years old.

**Tab 2.** Results of the antibodies survey: in the second column the number of people resulted positive to IgG (IgG+) antibody (29), in the third column the people positive to IgM antibody (IgM+), only 4 people show the presence of both antibodies (IgG+ and IgM+, third column), all IgM+ are also IgG+. In the last column it is shown people considered symptomatic according to the conditions described in the text: all were qPRC+ (that stands for qPRC-positive), IgG+ and IgM – (IgM-stands for IgM-negative).

| immunoassay 4/5 June 2020 |  |  |  |  |
| --- | --- | --- | --- | --- |
|  | number of participants | seroprevalence IgG | seroprevalence IgM | Symptomatic |
| OVERALL | 857 | 29 | 4<br>( IgG+) | 5<br>( qPCR+) (IgM-IgG+) |
| SEX |  |  |  |  |
| male | 393 | 13 | 3 | 3 |
| female | 464 | 16 | 1 | 2 |
| AGE-YEARS |  |  |  |  |
| 14-24 | 130 | 2 |  |  |
| 25-49 | 261 | 12 | 1 |  |
| 50-75 | 357 | 11 | 2 | 5 |
| >= 76 | 109 | 4 | 1 |  |

### 3.2 Second Wave

Europe, after a decline in detected infection cases in summer 2020, faced with the emergence of the Covid-19 second wave. At the end of September the number of people infected in Italy raised once again. Basically the reasons for such upsurge are to be found in the suppression of social distance rules during the summer and, when the second pandemic wave was apparent at the end of September and a second partial lockdown was adopted at the beginning of October, it had less stringent rules: the schools were kept open as well as the hotels; shops and restaurants had to respect only a limitation on the opening time and on the maximum number of people that could be together indoor (in the first lockdown these activities were kept all closed for three months), the number of services excluded because considered essential were extended. In Italy, following the rise of infection curve, which on October 7, 2020 reached more than 3000 infections in a day, Italian Government, between October 8 and November 5, issued new increasingly restrictive containment measures extended to the whole territory. In the second wave of infections, the Municipality of Cogne followed the Italian Government rules to limit the spread of the epidemic, but this time without any early lockdown as happened at the onset of the first wave. A second serological survey, to establish the longitudinal changes in antibodies level during a period of six months, was carried out on December 15 2020 just on the 29 individuals tested positives at the first serological test. (see Tab.3). Between June and December 2020 the seroprevalence among the 29 participants who presented antibodies at the first test has been reduced to 11 individuals. The antibodies persistence above the positive threshold (established by the chemiluminescent immunoassay) at 6 months from the first assay resulted around 40% of the tested population. We were able to establish for sure the dates of SARS-CoV-2 infections, certified by positive swabs, only for the 5 positives symptomatic cases. We have not highlighted correlation between presence and severity of symptoms and antibodies persistence. From the results reported in Tab.3 we could notice that 2 people (two males) out of the 5 proved symptomatic on March, 2020, were still positive on December 15 at the second serological survey. In particular of these two cases the one which proved positive to both IgM and IgG at December survey (see Tab.3) will be the topic of the next paragraph. From October 1 to December 15, 2020, were carried out on Cogne residents 87 nasopharyngeal swabs and founded 43 SARS-CoV-2 positive (prevalence 49%) with 35 symptomatic infections and 7 deaths (see Tab.4).

**Tab 3.** Results of December 15, 2020, immunoassay survey. Only the people positive to June 4-5 survey were tested again. The unique case of IgM+ (the patient reinfected) was also IgG+, 10 IgG+ out of 11 are IgM-(negative). Among the 5 people ascertained symptomatic (qPCR+ and IgG+ to the first assay), only 2 (male, one of whom is the reinfected patient) have retained IgG+ in this survey.

| immunoassay 15 december 2020 |  |  |  |
| --- | --- | --- | --- |
|  | number of participants | seroprevalence IgG | seroprevalence IgM |
| OVERALL | 29 | 11 | 1<br>( IgG+) |
| SEX |  |  |  |
| male | 15 | 9 |  |
| female | 14 | 2 |  |
| AGE-YEARS |  |  |  |
| 14-24 | 1 |  |  |
| 25-49 | 7 | 4 |  |
| 50-75 | 15 | 5<br>( 2 male among the 5<br>qPCR+ symptomatic) | 1 |
| >= 76 | 6 | 2 |  |

**Tab 4.** Results of the nasopharyngeal swabs taken from October 1 up to December 15, 2020. There is a clear increase of positive people, symptomatic people and deaths with respect to the first pandemic wave

| nasopharyngeal swabs from 1 October to 15 December 2020 |  |  |  |  |
| --- | --- | --- | --- | --- |
|  | number of participants | positive | symptomatic | died |
| **OVERALL** | 87 | 43 | 35 | 7 |
| **SEX** |  |  |  |  |
| **male** | 42 | 22 | 18 | 4 |
| **female** | 43 | 21 | 17 | 3 |
| **AGE-YEARS** |  |  |  |  |
| **0-24** | 22 | 5 | 1 | - |
| **25-49** | 17 | 6 | 4 | - |
| **50-75** | 35 | 20 | 18 | - |
| **>= 76** | 13 | 12 | 12 | 7 |

### 3.3 A case of secondary infection

Among the 11 individuals tested positives to the second assay (see Tab.3), we identified a case of reinfection. A 73-year-old male with a past medical story of cancer, on March 16, 2020 developed flu-like symptoms and was confirmed as SARS-CoV-2 positive case by qPCR on March 22. He did not required hospitalization and he has been successfully treated at home with dexamethasone 4 mg/d for 3 days. He experienced resolution of symptomatology in 4 days. On June 4, this patient, 80 days after the onset of symptoms, submitted to the serological assay by chemiluminescence organized by the Municipality, tested positive to SARS-CoV-2 with the following values: IgG = 34,19 AU/ml, IgM = 3,70 AU/ml (negative< 10). On 25 October he received pneumococcal and influence vaccine. On November 1st he had a close contact, within his own family, with a female presenting flu-like symptoms confirmed then SARS-CoV-2 positive by qPCR and on November 4, he developed complete ageusia, anosmia and flu-like symptoms. On November 5, he resulted positive to the nasopharyngeal swab and the 12-th day of worsening symptoms on November 16 he sought emergency care where chest X ray revealed a pattern of acute viral pneumonia typical of COVID-19. The pulmonary changes disappeared on the 13-th day after the hospitalization and the patient was released from the hospital (data from the hospital medical record, obtained with the written permission of the patient). On December 15, he repeated the Cogne’s Covid-19 serology survey, since positive to the first test of June 4, with the following result: IgG = 4000 AU/ ml, IgM= 19 AU/ml (negative<10).

## 4. Discussion

The current study attempts to asses the impact of the Sars -CoV-2 infection on the geographic, and arguably genetic isolated (as attested by the low number of different surnames compared with the number of families [6]) population of the Municipality of Cogne. The assay was performed during the course (March-December 2020) of the first and the second wave of SARS-CoV-2 infection in Italy. The spread of an infectious disease in a population is sustained by the transmissibility of the pathogen responsible for the infection and by the random contacts between individuals. The rate of spread is represented by the basic reproductive coefficient R0 that is determined by a combination of virus properties and contacts rate.

As general rule the impact of an infectious disease on a susceptible population is determined not only by the mobility patterns, but also by the structural, social and geographic features of the population and in the absence of pharmacological safeguards against the outbreaks of epidemics can be reduced essentially by community-transmission control measures. The conditions of relatively geographic isolation of the Municipality of Cogne, as well as very similar circumstances sometimes experienced in confined spaces of cruise ships (e.g. the Diamond Princess cruise ship [7]), provide the uncommon chance to study the spreading of infectious diseases within a community assumed to be fairly homogeneously mixed (any contact between individuals in a population occurs randomly and with equal probability) and restricted in a narrow and bounded area where it is easy to keep track of the infection dynamics.

As formerly suggested [8] the contact rate is dependent on population density: the contact rate tends to increase with density but rapidly saturate at higher densities. Compared to approximately 24000 people per Km^2^ on the Diamond Princess cruise ship and to 6000 people per Km^2^ in urban Wuhan, the urban density of Cogne’s population is 1080 people per Km^2^.

On March 4, 2020 the closure, by police control, of the only access route to Cogne (excluded essential business), has made the opportunity to prevent the introduction in Cogne of further infected cases from outside. Taken into account that the first symptomatic patient in Cogne dates to 13 March 2020, and given that the latency time of infection is 7/10 days, presumably the virus has entered the country around the first week of March just when the lockdown of the municipality started. This coincidence has supplied the rare chance to highlight the spread of an infection in an urban tissue, ruling out the contributions to the diffusion of the virus sustained by continuous new sources of infection coming from outside. The result of the countrywide lockdown, achieved through border-control measures, community-transmission control measures, and isolation of cases and their contacts, was striking: during the first wave Cogne counts 5 symptomatic patients only, all belonging to a single outbreak developed in the Municipal House, no one was hospitalized and no death. During the second pandemic wave it was not observed the same positive behavior, in fact the impact on the Cogne population was much worse. The differences between the first and second wave are the lockdown rules (much more strict during the first wave, from March 4 to May 18, 2020) and the timeliness of its enforcement (in fact the Cogne municipality advance the national lockdown on their territory). During the second wave the municipality followed the national Italian rules, without taking special initiative at local level. Along the second wave in the period from October 2020 up to the end of the year, around three months time, 7 deaths were counted (against zero). Between the deaths 100% were people over 80 years old, 29% showed only COVID-19 pathology without evidence of other diseases, 71% with preexisting medical conditions, especially heart disease, lung disease, diabetes or cancer and 86% lived in micro-community for elderly people (source: Cogne’s Municipality Administration). In the first wave all symptomatic people are concentrated in the age interval between 50 and 74 years old but with the second wave the eldest had to pay the highest cost to the pandemic and, as it happened in other Italian regions, the elderly shelters became a dangerous source of infection. From October 2020 up to the end of the year, 87 nasopharyngeal swabs were done, 43 resulted positive (49%), 35 symptomatic infections, against the 5 of the first wave, seven times more. Notably, nasopharyngeal swabs are tested for the presence of SARS-CoV-2 and can only detect active infection, not exposure but thanks to the cross-sectional serological survey we are confident to have clarified the infection rates of the Cogne population in the period under analysis [9]. Strategies to ease restraints on human mobility and interaction without provoking a major resurgence of transmission and mortality will depend on accurate estimates of population levels of infection and immunity. If we look at the car traffic [10] registered on the only road that connects Cogne to the bottom of the valley (the only possible motorway to Cogne) it is apparent that during the month of March 2020 an absolute minimum was registered (only around the 5% of usual seasonal traffic from Cogne and to Cogne was registered) with a small increment in April 2020 (about 10% of the seasonal traffic registered). The usual traffic conditions were reached for the summer (end of June, beginning of July, Cogne is a touristic place). From October up to December it was not any more observed such an impressive traffic decrement (only between 10% to 20% less). In the time interval under investigation (March 4, December 15, 2020) in Italy SARS-CoV-2 sensible mutations have not yet been spotted, there are proofs of their presence from the middle of January 2021 on, therefore the two pandemic waves could be associated with the same kind of virus, except for the mutation D614G in Spike protein, emerged in early 2020 and by June 2020 became the dominant form of the virus. On this topic a report from COVID-19 Genomics UK(COG-UK) Consortium [11] concluded that lineages bearing the 614G genotype grow ‘marginally’ faster than lineages bearing the original 614D genotype. The mathematics of the evolution of an epidemic are interestingly similar to those of a nuclear fission chain reaction (just by replacing neutrons with virus). According to our point of view, one of the principal lessons offered by the Chernobyl disaster, caused by an uncontrolled nuclear chain reaction, is that when a nuclear reaction get out of control is remarkably hard and highly expensive to contain it. A pandemic is a sort of biological Chernobyl, limiting the spread of the virus is similar to try of controlling a nuclear fission chain reaction. In order to slow the nuclear reaction control rods are inserted in the reactor to absorb neutrons, social distancing policies play for the virus the same role of rods for neutrons. A fission reaction releases in average between 2 to 3 neutrons, if more than one neutron is effective in inducing fission in another nucleus the reaction grows uncontrolled. Usually the fission reaction state is represented by the parameter K, defined as the ratio of the number of fissions produced in one step of neutron generation to the number of fissions in the preceding step. If the value of K is less than 1 the reaction turn off, if K=1 the reaction is sustained and if K is greater than 1 the reaction grows uncontrolled even up to an atomic explosion. By analogy the shutdown of an epidemic is generally reachable maintaining the rate of spread coefficient R0 below the threshold value of 1. To achieve this goal measures of social distancing and personal protection are introduced in the population to slow down the virus spread. Likewise the later the control rods are inserted in the nuclear reactor the less their action is going to be successful then the efficacy of the enforcement of severe lockdown in face of an advanced pandemic is timing-of-adoption dependent. SARS-CoV-2 is a respiratory virus, its transmission dynamics, unlike what happens with the neutron clouds for nuclear fission, still lacks of evidence. Analysis of 75,465 COVID-19 cases in China [12] has been cited as evidence that SARS-CoV-2 is primarily spread through direct person-to-person contact and respiratory droplet transmission. But the SARS-CoV-2 spreading process is more complicated because, as other studies [13] suggest, it may also be spread through indirect routes, such as contaminated surfaces and airborne transmission. In these cases, before SARS-CoV-2 can attach a new hosts, it must be released to the environment. Transmission from respiratory droplets occurs when a person touches a contaminated surface or gets caught directly in the spray zone of an infected patient. The 1 meter guidelines of social distancing originated from preliminary evidence that SARS-CoV-2 is mainly spread through respiratory droplet transmission and that heavy droplets do not travel in average to more than 1 meter before landing. This approach could remind the free mean neutron path of the fission reactions, but without the same highly precise measures. How the expelled droplets impact disease transmission depends on the number of droplets produced, size of the droplets and concentration of pathogenic hitchhikers on board. van Doremalen et al. [14] recently reported that SARS-CoV-2 remained viable in aerosols for 3 hours in a laboratory setting (this figure is equivalent to the mean lifetime for the neutrons in the fission reactions) but as far as we know, nothing is known about outside the lab. Recommended non-pharmacological interventions to constraint SARS-CoV-2 spread include avoiding close contact with people above all who have active symptoms, wearing a face mask and practicing good hand hygiene. It must be stressed that the spread of airborne viruses is more difficult to control than the spread of viruses through person-to-person contact alone. Therefore it is not only important to avoid crowded closed room but also do not enter highly frequented room even when it is almost empty and this case could apply to restaurants, pubs and shops, as the difference between Cogne’s collected data of the first and second wave seem to suggest according to the different rules of social distance that were applied in the two cases (in the second wave the shops and pubs were kept open with only two people indoor at the time, restaurants were open with reduced number of customers inside). The distance that airborne particles can travel and the length of time they remain suspended in the air is much greater. Therefore, measures of strong social distancing policies are much more important with airborne viruses as the data of this study may suggest. Another outcome of the present study claims that the antibodies persistence above the positive threshold at 6 months from the first assay resulted around 40% of the tested population. This result suggests that antibodies anti SARS-CoV-2 decay faster than other virus such as measles, rubella and mumps [15] (unlike humoral immunity memory T cells persist for much more time [16]). From present investigation only one case of secondary infection has been observed and up to now secondary infection with SARS-CoV-2 has been reported in very few cases worldwide [17], [18], [19] and it is still poorly understood [20]. In the case of COVID-19 recurrence here presented, the patient experienced two symptomatic episodes within six months of each. Unfortunately for this case it was not available the genomic sequencing of the virus. The arguments that could be adduced to explain the observed second more severe infection are merely speculative and at the root of the severe relapse of the patient analyzed we suggest the following three pathophysiological hypotheses:

a. viral reinfection due to new prolonged contact with a SARS-CoV-2 infected member within the household. Given that the recurrence occurred in the same relatively isolated municipality, the effective possibility of a new variant of the virus at work is low (except for the D614G mutation), therefore the present hypotheses is likely attributable to the lack (e.g. by early antibodies decay) of protective immunity able to prevent relapse or attenuate symptoms due to reinfection. Anyway, at first instance it is strongly conceivable that the severity of symptoms observed in the second infection was basically due to a particular high viral load.
b. viral reactivation due to a suboptimal control of the first infectious episode thus enabling a later reactivation of the same virus. This hypotheses is strengthened by the fact that the patient cured the first episode of SARS-CoV-2 infection with a corticosteroid, an immunosuppressive drug, that may have determined the failure of virus eradication and subsequent reactivation.
c. dysregulated and aberrant immune response. The development of acute distress respiratory syndrome (ARDS) in the case here presented coincides, as usual, with appearance of the antibodies arising from the activation of the adaptive immune response suggesting a role of humoral immunity in the implementation of the inflammatory rebound. Antibody-Dependent Enhancement (ADE) [21] is a little known and potentially dangerous phenomenon where the binding of a virus by non neutralizing antibodies, produced by previous exposure of virus, may paradoxically worsen rather than suppress an infection by enhancing viral uptake and replication. The mechanism of ADE is experimentally well documented (e. g. in dengue reinfection [22]) and actually strongly suspected to be implicated in the enhanced disease severity in SARS-CoV-2 re-infected patients [23].

## 5. Conclusions

Here we summarize the conclusions deduced from the present study on the SARS-CoV-2 impact on the Municipality of Cogne. Firstly, we collect enough evidence to claim that the containment of the infection spread is possible through the stringent and, most important, early lockdown. The application of early and severe social distancing measures is currently and arguably the most powerful instrument to contain the pandemic spread, allowing the achievement of two urgent and important goals: the improvement of the powerful tool of contact tracing and quarantine and the consequent lightening of the load on critical care resources. Delayed lockdown approach has shown far weaker effectiveness in preserving the same population from the widespread outbreak of SARS-CoV-2. The effectiveness of severe lockdown to curb the spread of the virus, in a specific context as that in the present study, has shown strong dependence to the timing of the adoption of the restrictions. We could not exclude that the same topics stay equally valid if extended to larger scale populations as suggested by previous research [24],[25]. Secondly, our findings, in the observational period of six months, regarding the decay of humoral immunity in about 60% of the tested population, independently of the severity of symptomatology (with one case of reinfection), raise concern about the long-term maintenance of humoral immunity against SARS-CoV-2. The identified case of a second COVID-19 infection with remarkably worse symptoms as compared with the first bout, focus attention on the potential ability of SARS-CoV-2 to elude acquired immunity and on the contingency of rogue immune responses that could seriously damage tissues and organs. One of the key features in the achievement of herd immunity is the persistence in time of the neutralizing antibodies titer against the pathogen in the population.

Moreover, as highlighted in the present work, short term immunity along with the possibility of reinfection complicate the long-lasting vaccine efficacy. In such a challenging scenario, delayed and intermittent distancing measures could favor the waving mechanism of resurgence, in agreement with the present data and with the data of the 1918 flu pandemic [26], jeopardizing the necessity of a rapid achievement of population immunity. The present study aimed to provide sound scientific evidence as a basis for informed policy direction and intervention for constraint the infection.

## Data Availability

All data can be requested at

## Competing interests

Both authors have no conflict of interest to disclose.

## Authors contribution

Both authors conceptualized the study. F.T. designed the experiments, both authors analyzed the data, wrote the manuscript, reviewed and edited the manuscript.

## Acknowledgments

The authors gratefully acknowledge all the participants to the study, the mayor Franco Allera, the Municipal Council and the Municipal Executive, the administrative personnel and health-care workers who collaborated on this study and especially Maria Grazia Brunero, Rinaldo Billia, Andrea Celesia, Giuseppe Cutano, Massimo Martinengo, Piergiorgio Montanera, Claudio Perratone, Piero Roullet, Sofia Truc and Paolo Varone.

## Notes

### Competing Interest Statement

The authors have declared no competing interest.

### Funding Statement

Research budgeted by Municipality of Cogne under the contract COGNE_COVID

### Author Declarations

IRB: Ethics Committee of Aosta Valley (I).

## Reference

1. http://tipografiavaldostana.com/it/editoria/il-messager-valdotain, “Le Messager Valdôtain”, 1919, xpp. 49.

2. Whitman, J.D. et al., Evaluation of SARS-CoV-2 serology assays reveals a range of test performance. Nature Biotechnology 38(10), 2020, pp. 1174–1183,https://doi.org/10.1038/s41587-020-0659-0

3. Zhao, J. et al., Antibody responses to SARS-CoV-2 in patients of novel coronavirus disease. Clin. Infect. Dis., 71(16), 2020, pp. 2027–2034, https://doi.org/10.1093/cid/ciaa344 (2020).

4. Weber, M.C. et al., Characteristics of three different Chemiluminescence Assays for testing for SARS-CoV-2 antibodies. Disease Markers (2021), 2021, Article number 8810196, doi: 10.1155/2021/8810196

5. Barra, G.B. et al., Analytical Sensitivity and Specificity of Two RT-qPCR Protocols for SARS-CoV-2 Detection Performed in an Automated Workflow. Genes 11, 2020, pp. 1183–1197; doi:10.3390/genes11101183

6. Fiorito, G. et al., The Italian genome reflects the history of Europe and the Mediterranean basin. Eur. J. Hum. Genet. 24, 2016, pp. 1056–1062, doi:10.1038/ejhg.2015.233.

7. Rocklov, J. et al., COVID-19 outbreak on the Diamond Princess cruise ship: estimating the\ epidemic potential and effectiveness of public health countermeasures. Journal of travel Medicine 27(3),2020, pp1–7, doi: 10.1093/jtm/taaa030

8. Hu, H., Nigmatulina, K., Eckhoff, P. The scaling of contact rates with population density for the infectious disease models. Mathematical Bioscience 244(2), 2013, pp.125–34, doi: 10.1016/j.mbs.2013.04.013

9. Lavezzo, E. et al., Suppression of a SARS-CoV-2 outbreak in the Italian municipality of Vo’. Nature 584, 2020, pp. 425–429, https://doi.org/10.1038/s41586-020-2488-1

10. EnelX & HERE Technologies, City Analytics-Mappa di mobilità, https://www.enelx.com/it/ it/smart-city/soluzioni/soluzioni-smart-city/city-analytics/dashboard-covid-19

11. Report #9 - 25th June 2020 - COVID-19 Genomics UK (COG-UK) Consortium https://www.cogconsortium.uk/wp-content/uploads/2020/07/25th-June-2020-Report-COVID-19-Genomics-UK-COG-UK-Consortium.pdf (accessed August 15, 2020)

12. Sean Wei Xiang Ong et al., Air, Surface Environmental, and Personal Protective Equipment Contamination by Severe Acute Respiratory Syndrome Coronavirus 2 (SARS-CoV-2) From a Symptomatic Patient. JAMA. 323(16), 2020, pp. 1610–1612; doi:10.1001/jama.2020.3227

13. Morowska, L. et al., How can airborne transmission of COVID-19 indoors be minimised? Environment International 142, 2020, 105832, https://doi.org/10.1016/j.envint.2020.105832

14. van Doremalen, N. et al., Aerosol and Surface Stability of SARS-CoV-2 as Compared with SARS-CoV-1. N. Engl. J. Med. 382, 2020, pp. 1564–1567; doi: 10.1056/NEJMc2004973.

15. Seow, J. et al., Longitudinal observation and decline of neutralizing antibody responses in the three months following SARS-CoV-2 infection in humans. Nat. Microbiol. 5, 2020, pp. 1598– 1607, https://doi.org/10.1038/s41564-020-00813-8

16. Zuo, J. et al., Robust SARS-CoV-2-specific T-cell immunity is maintained at 6 months following primary infection. bioRxiv [Preprint] (2020). https://doi.org/10.1101/2020. 11.01.362319 (accessed 2 November 2020).

17. Tillett, R.L. et al., Genomic evidence for reinfection with SARS-CoV-2: a case study. Lancet Infect Dis. Published online October 12, 2020. doi:10.1016/S1473-3099(20)30764-7

18. To, K.K-W. et al., COVID 19 re-infection by a phylogenetically distinct SARS-coronavirus-2 strain confirmed by whole genome sequencing. Clin Infect Dis. Published online August 25, 2020. doi:10.1093/cid/ciaa1275

19. Loconsole, D. et al., Recurrence of COVID-19 after recovery: a case report from Italy. Infection 2020, pp.1–3 [published online ahead of print, 2020 May 16]. doi:10.1007/s15010-020-01444-1

20. Iwasaki, A. What reinfections mean for COVID-19. Lancet Infect. Dis. 21(1), 2021, pp. 3–5, https://doi.org/10.1016/S1473-3099(20)30783-0 (2020).

21. Iwasaki, A. and Yang, Y., The potential danger of suboptimal antibody responses in COVID-19. Nat. Rev. Immunol. 20, 2020, pp. 339–341, https://doi.org/10.1038/s41577-020-0321-6

22. Dejnirattisai, W., et al., Cross-reacting antibodies enhance dengue virus infection in humans. Science 328(5979), 2010, pp.745–748, doi: 10.1126/science.1185181.

23. Ulrich, H., Pillat, M., & Tarnok, M., A. Dengue Fever, COVID-19 (SARS-CoV-2), and Antibody-Dependent Enhancement (ADE): A Perspective. Cytometry A, 97(7), 2020, pp. 662–667, doi: 10.1002/cyto.a.24047.

24. Baker, M.G., Kvalsvig, A., Verrall, A.J., Wellington, N. New Zealand’s COVID-19 elimination strategy. Med. J. Aust. 213, 2020, pp.198–200, doi: 10.5694/mja2.50735

25. Summers, D.J., Cheng, D. H.-Y., Lin, P.H.-H., Barnard, D.L.T., Kvalsvig, D.A., Wilson, P.N., Baker, P.M.G. Potential lessons from the Taiwan and New Zealand health responses to the COVID-19 pandemic. Lancet Reg. Health West. Pac. 4, 2020, 100044. doi:10.1016/j.lanwpc.2020.100044

26. Hatchett R.J., Mecher, C.E., Lipsitch M. Public health interventions and epidemic intensity during the 1918 influenza pandemic. Proc. Natl. Acad. Sci. U.S.A. 104, 7582–7587 (2007). doi:10.1073/pnas.0610941104pmid:17416679

